# REACT-1 round 7 interim report: fall in prevalence of swab-positivity in England during national lockdown

**DOI:** 10.1101/2020.11.30.20239806

**Authors:** Steven Riley, Oliver Eales, Caroline E. Walters, Haowei Wang, Kylie E. C. Ainslie, Christina Atchison, Claudio Fronterre, Peter J. Diggle, Deborah Ashby, Christl A. Donnelly, Graham Cooke, Wendy Barclay, Helen Ward, Ara Darzi, Paul Elliott

## Abstract

**Background:** The second wave of the 2020 COVID-19 pandemic in England has been characterized by high growth and prevalence in the North with lower prevalence in the South. High prevalence was first observed at younger adult ages before spreading out to school-aged children and older adults. Local tiered interventions were in place up to 5th November 2020 at which time a second national lockdown was implemented.

**Methods:** REACT-1 is a repeated cross-sectional survey of SARS-CoV-2 swab-positivity in random samples of the population of England. The current period of data collection (round 7) commenced on 13th November 2020 and we report interim results here for swabs collected up to and including 24th November 2020. Because there were two distinct periods of growth during the previous round 6, here we compare results from round 7 (mainly) with the second half of round 6, which obtained swabs between 26th October and 2nd November 2020. We report prevalence both unweighted and reweighted to be representative of the population of England. We describe trends in unweighted prevalence with daily growth rates, doubling times, reproduction numbers (R) and splines. We estimated odds ratios for swab-positivity using mutually-adjusted multivariable logistic regression models.

**Results:** We found 821 positives from 105,123 swabs giving an unweighted prevalence of 0.78% (95% CI, 0.73%, 0.84%) and a weighted prevalence of 0.96% (0.87%, 1.05%). The weighted prevalence estimate was ∼30% lower than that of 1.32% (1.20%, 1.45%) obtained in the second half of round 6. This decrease corresponds to a halving time of 37 (30, 47) days and an R number of 0.88 (0.86, 0.91). Using only data from the most recent period, we estimate an R number of 0.71 (0.54, 0.90). A spline fit to prevalence showed a rise shortly after the previous period of data collection followed by a fall coinciding with the start of lockdown. The national trends were driven mainly by reductions in higher-prevalence northern regions, with prevalence approximately unchanged in the Midlands and London, and smaller reductions in southern lower-prevalence regions. Sub-regional analyses showed variable changes in prevalence at the local level including marked declines in the North, but also local areas of growth in East and West Midlands. Mutually adjusted models in the most recent period indicated: people of Asian ethnicity, those living in the most deprived neighbourhoods, and those living in the largest households, had higher odds of swab-positivity.

**Conclusion:** Three weeks into the second national lockdown in England there has been a ∼30% proportionate reduction in prevalence overall, with greater reductions in the North. As a result, inter-regional heterogeneity has reduced, although average absolute prevalence remains high at ∼1%. Continued monitoring of the epidemic in the community remains essential until prevalence is reliably suppressed to much lower levels, for example, through widespread vaccination.

## Introduction

England entered a second national lockdown in response to the COVID-19 pandemic on 5th November 2020. This followed rises in prevalence across the country, and high prevalence of people testing positive for SARS-CoV-2 virus especially in the North [1]. The REal-time Assessment of Community Transmission-1 (REACT-1) study has been undertaking surveillance of the epidemic in England since May 2020 [2] using self-administered throat and nose swabs approximately monthly among random samples of the general population. We recently reported results from the sixth round of data collection, which was undertaken in two halves, from 16th to 25th October [3] and from 26th October to 2nd November 2020 [1]. In the most recent round 6 data (second half) we found evidence for a fall then rise in prevalence. Here we present interim round 7 results, denoted 7a, covering the first period of data collection in this round (13th - 24th November 2020).

## Methods

REACT-1 methods are published [2]. Briefly, we send a letter of invitation to named individuals randomly selected from the list of GP patients held by the National Health Service (NHS), stratified by lower-tier local authorities (LTLAs, n=315) in England. Following registration, we obtain self-administered throat and nose swabs (or swabs administered by parent/guardian for children ages 5 to 12 years) which are then sent on a cold chain for analysis by RT-PCR in a single laboratory. Participants are also invited to complete a brief questionnaire. Achieved sample sizes have ranged from 120,000 to 175,000 across the six rounds of data collection from May to beginning of November 2020 [1,2].

We estimate prevalence of SARS-CoV-2 infection (and 95% confidence intervals) by age, sex, region and other socio-demographic and clinical characteristics both unweighted and weighted to be representative of the population of England. For each round of data collection, we estimate time-trends of prevalence of people testing positive both between successive rounds and within rounds. We use exponential growth and decay models to describe the trends and multivariable logistic regression to investigate associations of key covariates with odds of testing positive on RT-PCR. We also assess sub-regional trends in SARS-CoV-2 prevalence using a neighbourhood prevalence statistic as previously described [3].

All statistical analyses are carried out in the R environment [4].

Research ethics approval has been obtained from the South Central-Berkshire B Research Ethics Committee (IRAS ID: 283787).

## Results

In this first part of round 7 (round 7a) we found 821 positives from 105,123 swabs giving an unweighted prevalence of 0.78% (95% CI, 0.73%, 0.84%) and a weighted prevalence of 0.96% (0.87%, 1.05%). The weighted prevalence estimate was ∼30% lower than that of 1.32% (1.20%, 1.45%) obtained in round 6b, the second half of round 6 (Table 1), and has now returned to levels seen in mid-October (Figure 1, Figure 2).

**Table 1.** Unweighted and weighted prevalence of swab-positivity across seven rounds of REACT-1.

| Round | Tested swabs | Positive swabs | Unweighted prevalence (95% CI) | Weighted prevalence (95% CI) | First sample | Last sample |
| --- | --- | --- | --- | --- | --- | --- |
| 1 | 120,620 | 159 | 0.13% (0.11%, 0.15%) | 0.16% (0.13%, 0.19%) | 1/5/2020 | 1/6/2020 |
| 2 | 159,199 | 123 | 0.077% (0.065%, 0.092%) | 0.088% (0.068%, 0.11%) | 19/6/2020 | 7/7/2020 |
| 3 | 162,821 | 54 | 0.033% (0.025%, 0.043%) | 0.040% (0.027%, 0.053%) | 24/7/2020 | 11/8/2020 |
| 4 | 154,325 | 137 | 0.089% (0.075%, 0.11%) | 0.13% (0.096%, 0.15%) | 20/8/2020 | 8/9/2020 |
| 5 | 174,949 | 824 | 0.47% (0.44%, 0.50%) | 0.60% (0.55%, 0.71%) | 18/9/2020 | 5/10/2020 |
| 6 | 160,175 | 1732 | 1.08% (1.03%, 1.13%) | 1.30% (1.21%, 1.39%) | 16/10/2020 | 2/11/2020 |
| 6a | 85,965 | 863 | 1.00% (0.94%, 1.07%) | 1.28% (1.16%, 1.42%) | 16/10/2020 | 25/10/2020 |
| 6b | 74,210 | 869 | 1.17% (1.10%, 1.25%) | 1.32% (1.20%, 1.45%) | 26/10/2020* | 2/11/2020 |
| 7a | 105,123 | 821 | 0.78% (0.73%, 0.84%) | 0.96% (0.87%, 1.05%) | 13/11/2020** | 24/11/2020** |

\*Includes small number of samples from prior period
\*\*Small number of samples collected on 13 - 15 Nov and 24 Nov
Round 7a is updated with data on 25 Nov, not complete.

**Figure 1.**
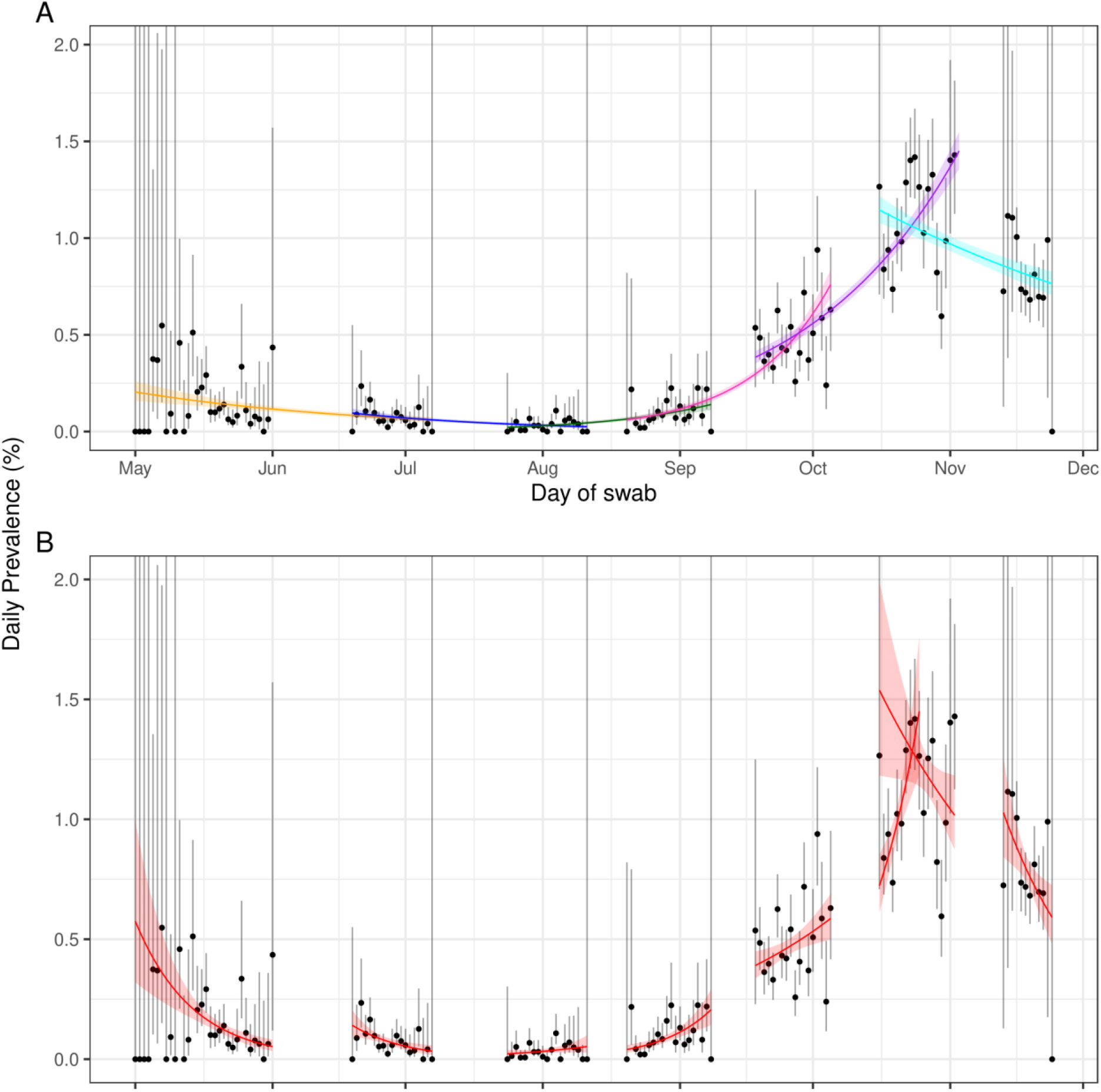
Constant growth rate models fit to REACT-1 data for sequential and individual rounds. **A** models fit to REACT-1 data for sequential rounds; 1 and 2 (yellow), 2 and 3 (blue), 3 and 4 (green), 4 and 5 (pink), 5 and 6 (purple), and 6 and 7a (cyan). **B** models fit to individual rounds 1, 2, 3, 4, 5, 6a, 6b and 7a (red). Vertical lines show 95% confidence intervals for observed prevalence (black points). Shaded regions show 95% posterior credibility intervals for growth models.

**Figure 2.**
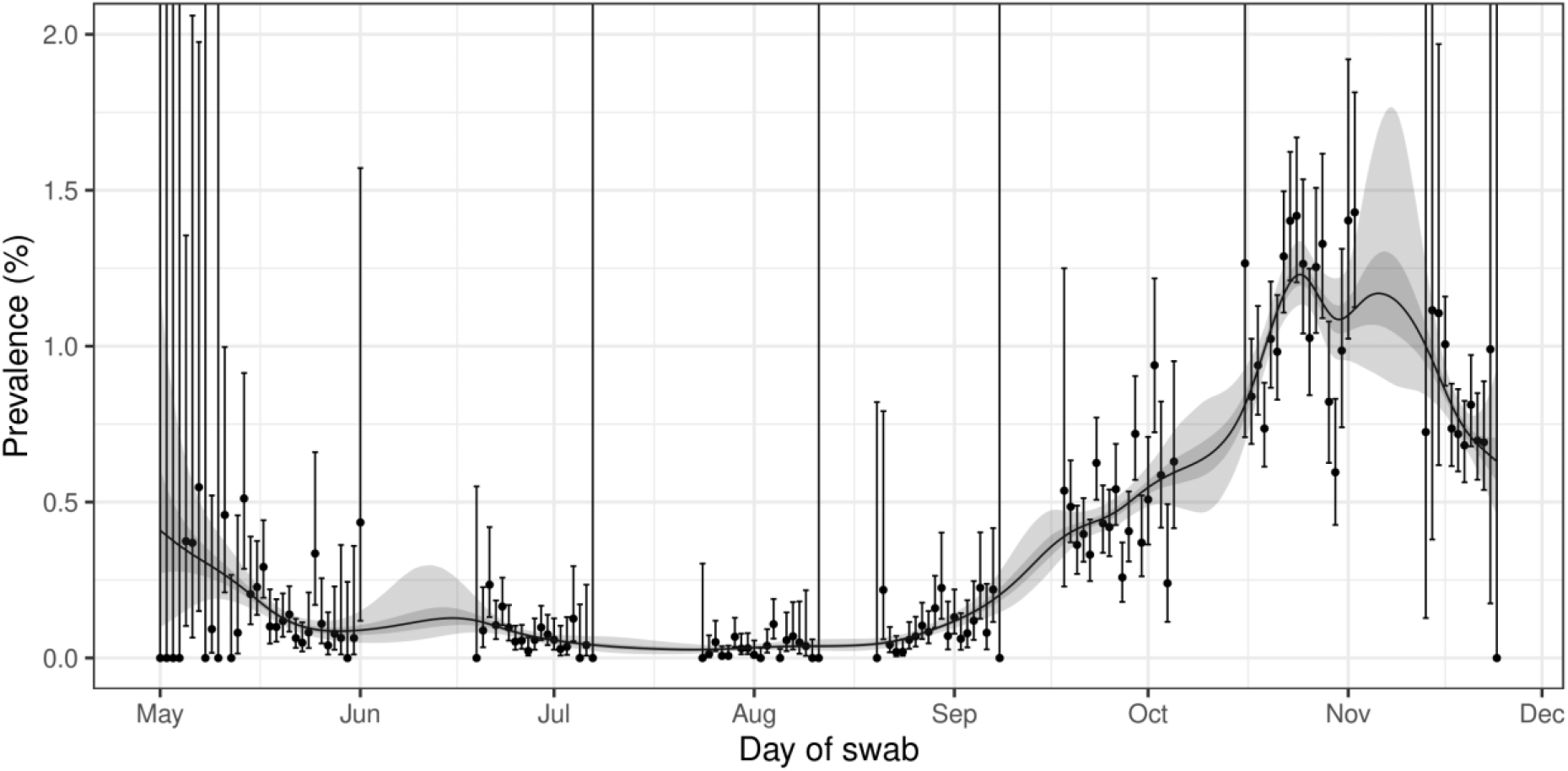
Prevalence of swab-positivity estimated using a p-spline for the full period of the study with central 50% and 95% credible intervals.

The decrease in prevalence between rounds 6b and 7a represents a national halving time of 37 (30, 47) days with a corresponding R number of 0.88 (0.86, 0.91) (Table 2, Figure 1). Using data only from within round 7a, we estimate an R number of 0.71 (0.54, 0.90) (Table 2, Figure 1). After the fall and rise previously reported for 6b [1], our spline estimate of prevalence suggests an increase into the start of the second lockdown followed by a decrease (Figure 2). Sensitivity analyses showed slightly lower R numbers for: double gene target positives, positives with a lower CT value cut-off for the N gene, and non-symptomatic participants (Table 2).

**Table 2.** National estimates of growth rate, doubling time and reproduction number for rounds 6b and 7a together, rounds 6 and 7a together, and for round 7a alone.

| Round | Outcome | Growth Rate | R | Probability R>1 | Doubling(+)/Halving(-) Time |
| --- | --- | --- | --- | --- | --- |
| 6b&7a | All positives** | -0.019 ( -0.023 , -0.015 ) | 0.88 ( 0.86 , 0.91 ) | <0.01 | -36.7 ( -30.1 , -47 ) |
|  | Non-symptomatics | -0.031 ( -0.037 , -0.024 ) | 0.82 ( 0.78 , 0.85 ) | <0.01 | -22.6 ( -18.6 , -29.1 ) |
|  | Positive for both E and N genes | -0.017 ( -0.022 , -0.012 ) | 0.90 ( 0.87 , 0.93 ) | <0.01 | -40.9 ( -31.6 , * ) |
|  | Positive for both E and N genes or positive only for N gene with CT 35 or less | -0.017 ( -0.021 , -0.012 ) | 0.90 ( 0.87 , 0.92 ) | <0.01 | -41.3 ( -32.6 , * ) |
| 6&7a | All positives** | -0.010 ( -0.013 , -0.007 ) | 0.94 ( 0.92 , 0.95 ) | <0.01 | * ( * , * ) |
|  | Non-symptomatics | -0.021 ( -0.026 , -0.016 ) | 0.87 ( 0.84 , 0.90 ) | <0.01 | -33.1 ( -26.9 , -42.7 ) |
|  | Positive for both E and N genes | -0.008 ( -0.012 , -0.004 ) | 0.95 ( 0.93 , 0.97 ) | <0.01 | * ( * , * ) |
|  | Positive for both E and N genes or positive only for N gene with CT 35 or less | -0.009 ( -0.012 , -0.006 ) | 0.94 ( 0.92 , 0.96 ) | <0.01 | * ( * , * ) |
| 7a | All positives** | -0.050 ( -0.085 , -0.016 ) | 0.71 ( 0.54 , 0.90 ) | <0.01 | -13.8 ( -8.2 , -43.2 ) |
|  | Non-symptomatics | -0.084 ( -0.144 , -0.024 ) | 0.55 ( 0.31 , 0.85 ) | <0.01 | -8.3 ( -4.8 , -28.4 ) |
|  | Positive for both E and N genes | -0.088 ( -0.129 , -0.046 ) | 0.53 ( 0.36 , 0.73 ) | <0.01 | -7.9 ( -5.4 , -15.1 ) |
|  | Positive for both E and N genes or positive only for N gene with CT 35 or less | -0.062 ( -0.100 , -0.026 ) | 0.65 ( 0.47 , 0.84 ) | <0.01 | -11.1 ( -6.9 , -27 ) |

\* Doubling/Halving time had an estimated magnitude greater than 50 days and so represented approximately constant prevalence
\*\* Positive for both E and N genes or positive only for N gene with CT 37 or less.

Between rounds 6b and 7a, there was a marked decrease in swab-positivity in the North West and North East, where there was a reduction in prevalence of over 50% (Table 3b, Figure 3). In contrast there was little change in prevalence between rounds 6b and 7a in East and West Midlands and London (Table 3b, Figure 3). The highest prevalence was in the West Midlands at 1.55% (1.14%, 2.10%) (Table 3b, Figure 3).

**Table 3a.**
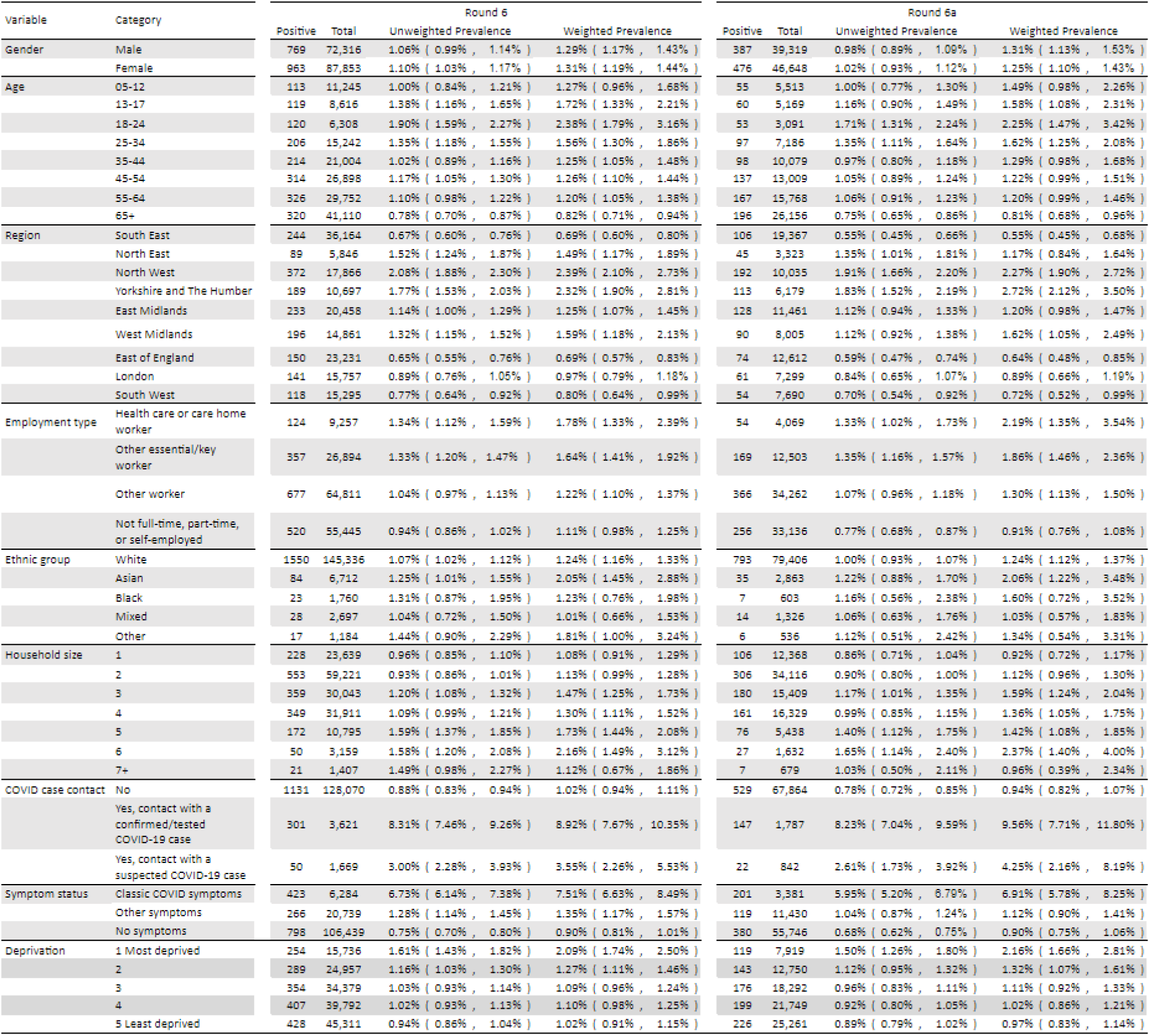
Unweighted and weighted prevalence of swab-positivity by variable and category for rounds 6 and 6a.

**Table 3b.**
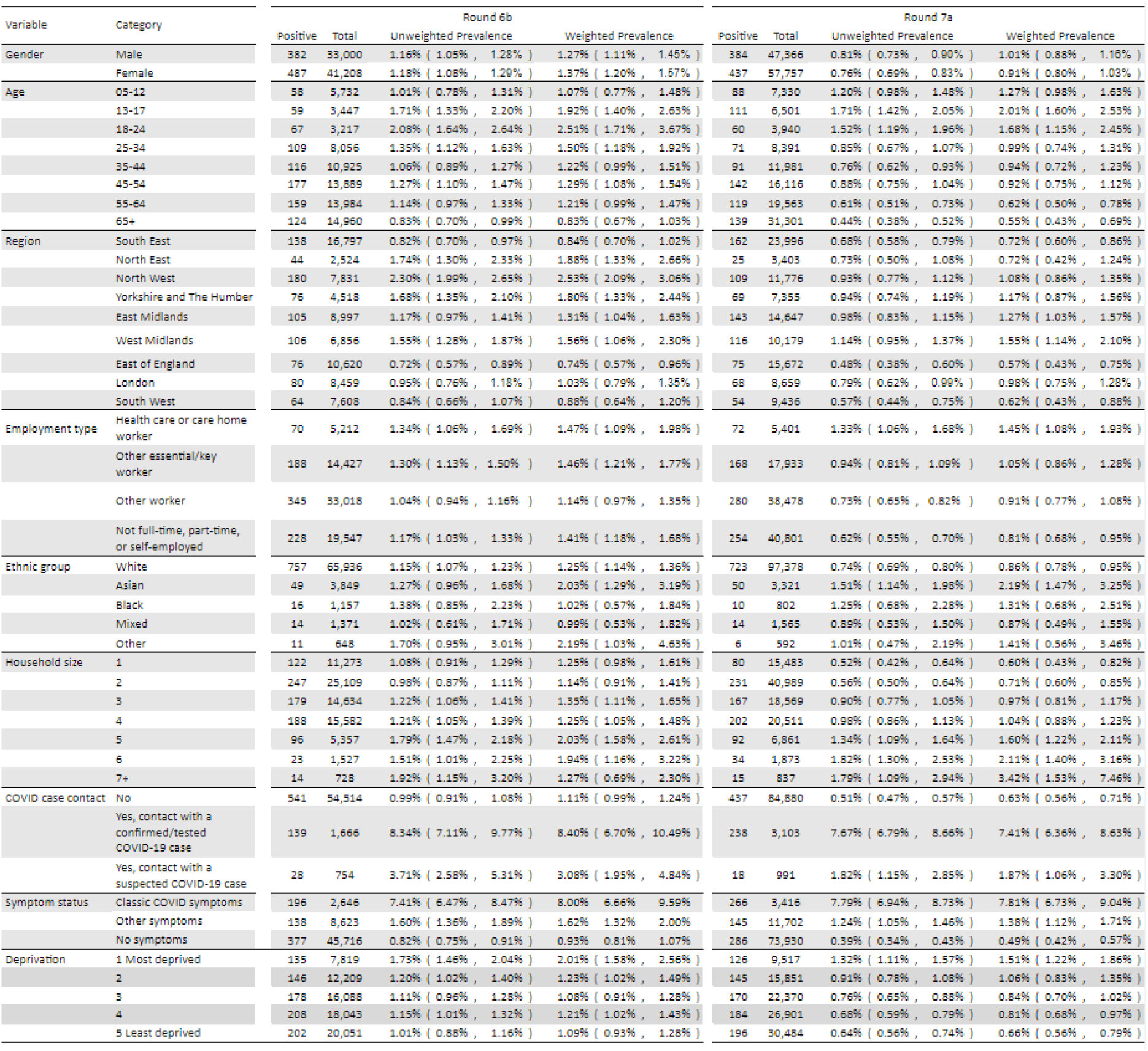
Unweighted and weighted prevalence of swab-positivity by variable and category for rounds 6b and 7a.

**Figure 3.**
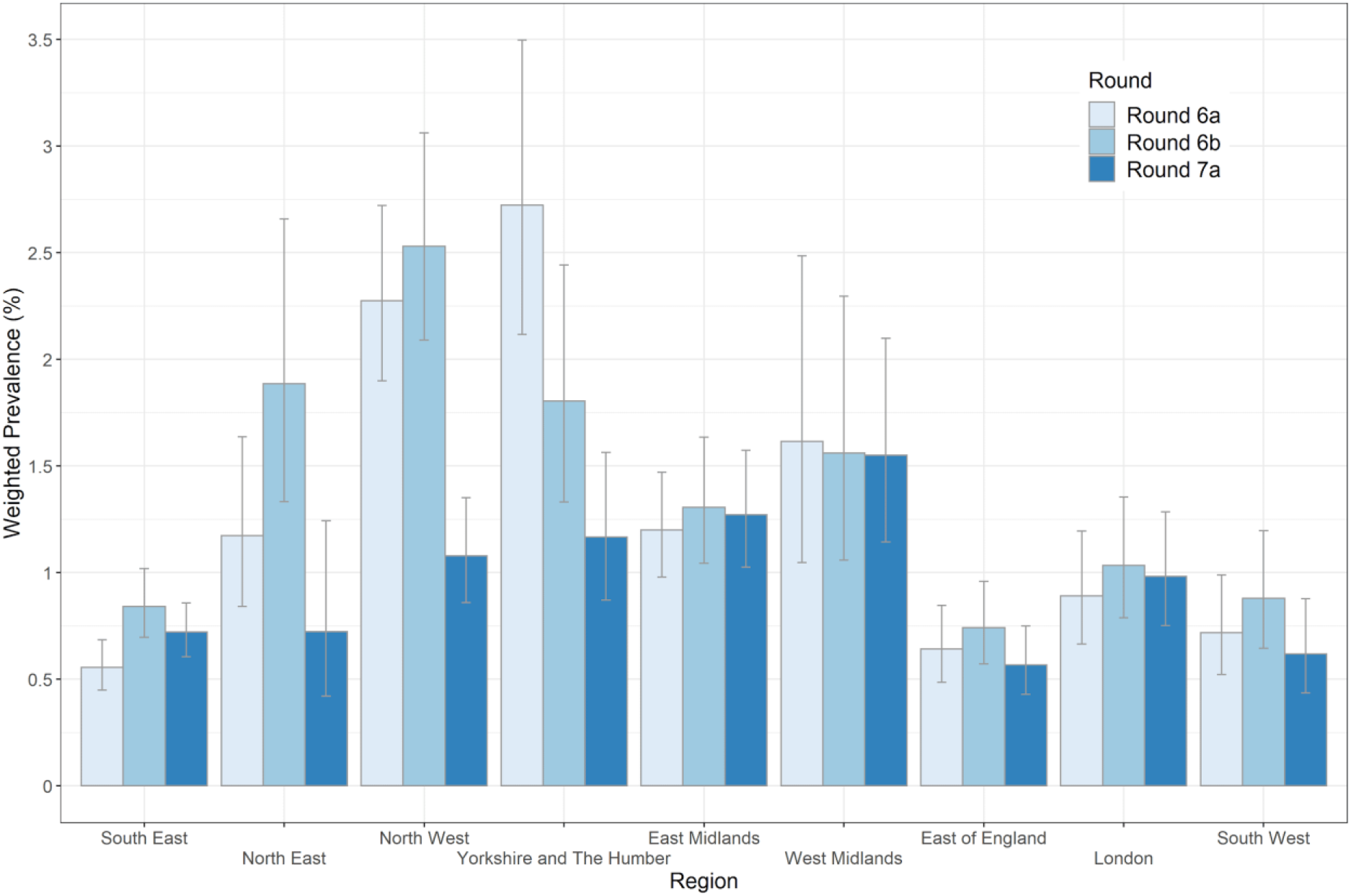
Weighted prevalence of swab positivity by region for rounds 6a, 6b and 7a. Bars show 95% confidence intervals.

At the regional level, R numbers between rounds 6b and 7a ranged from 0.76 (0.71, 0.82) for North West to 0.95 (0.86, 1.04) for London (Table 4). We also calculated regional average R numbers across the entire period of round 6 and 7a, resulting in similar, but slightly higher estimates (Table 4, Figure 4).

**Table 4.** Regional estimates of growth rate, doubling time and reproduction number for rounds 6b and 7a together, and rounds 6 and 7a together.

| Round | Region | Growth Rate | R | Probability R>1 | Doubling(+)/Halving(-) Time |
| --- | --- | --- | --- | --- | --- |
| 6b&7a | East Midlands | -0.010 ( -0.021 , 0.001 ) | 0.94 ( 0.87 , 1.00 ) | 0.03 | * ( -33.1 , * ) |
|  | West Midlands | -0.016 ( -0.027 , -0.005 ) | 0.90 ( 0.84 , 0.97 ) | <0.01 | -44.0 ( -25.5 , * ) |
|  | East of England | -0.018 ( -0.032 , -0.004 ) | 0.89 ( 0.81 , 0.97 ) | <0.01 | -38.6 ( -21.8 , * ) |
|  | London | -0.008 ( -0.022 , 0.006 ) | 0.95 ( 0.86 , 1.04 ) | 0.13 | * ( -31.1 , * ) |
|  | North West | -0.040 ( -0.050 , -0.030 ) | 0.76 ( 0.71 , 0.82 ) | <0.01 | -17.2 ( -13.8 , -23.0 ) |
|  | North East | -0.036 ( -0.057 , -0.016 ) | 0.78 ( 0.67 , 0.90 ) | <0.01 | -19.0 ( -12.1 , -43.8 ) |
|  | South East | -0.010 ( -0.020 , 0.000 ) | 0.94 ( 0.88 , 1.00 ) | 0.03 | * ( * , * ) |
|  | South West | -0.017 ( -0.033 , 0.000 ) | 0.90 ( 0.80 , 1.00 ) | 0.02 | -41.6 ( -21.2 , * ) |
|  | Yorkshire | -0.026 ( -0.040 , -0.012 ) | 0.84 ( 0.76 , 0.92 ) | <0.01 | -26.7 ( -17.2 , * ) |
| 6&7a | East Midlands | -0.006 ( -0.013 , 0.002 ) | 0.96 ( 0.92 , 1.01 ) | 0.06 | * ( * , * ) |
|  | West Midlands | -0.004 ( -0.012 , 0.004 ) | 0.97 ( 0.92 , 1.03 ) | 0.17 | * ( * , * ) |
|  | East of England | -0.008 ( -0.018 , 0.001 ) | 0.95 ( 0.89 , 1.01 ) | 0.05 | * ( -37.7 , * ) |
|  | London | -0.003 ( -0.013 , 0.008 ) | 0.98 ( 0.92 , 1.05 ) | 0.32 | * ( * , * ) |
|  | North West | -0.026 ( -0.033 , -0.019 ) | 0.84 ( 0.80 , 0.89 ) | <0.01 | -26.8 ( -20.8 , -37.1 ) |
|  | North East | -0.022 ( -0.037 , -0.008 ) | 0.86 ( 0.78 , 0.95 ) | <0.01 | -31.2 ( -18.6 , * ) |
|  | South East | 0.003 ( -0.005 , 0.010 ) | 1.02 ( 0.97 , 1.06 ) | 0.76 | * ( * , * ) |
|  | South West | -0.008 ( -0.020 , 0.003 ) | 0.95 ( 0.88 , 1.02 ) | 0.09 | * ( -35.0 , * ) |
|  | Yorkshire | -0.022 ( -0.032 , -0.013 ) | 0.87 ( 0.81 , 0.92 ) | <0.01 | -31.4 ( -21.7 , * ) |

\* Doubling/Halving time had an estimated magnitude greater than 50 days and so represented approximately constant prevalence

**Figure 4.**
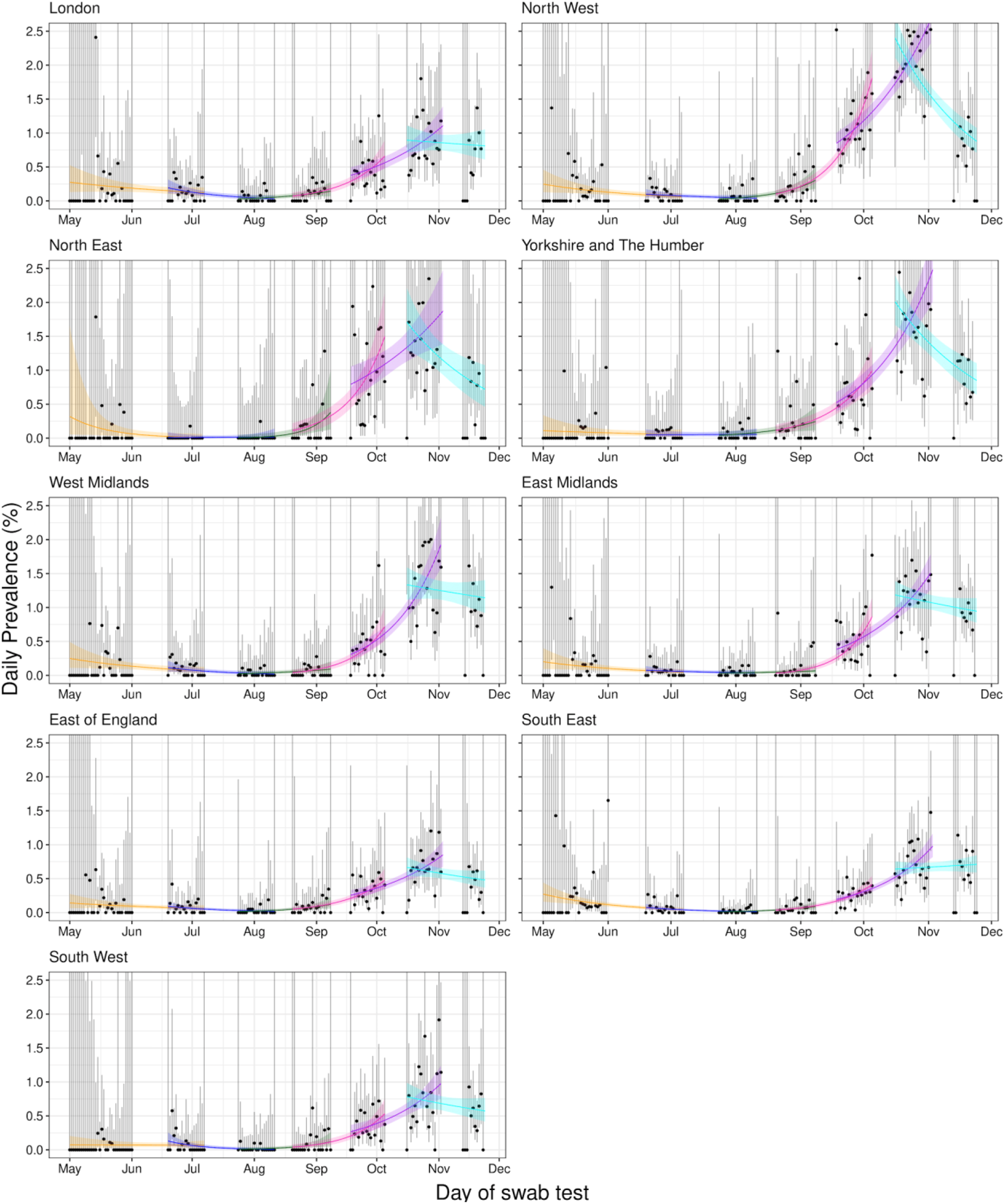
Constant growth rate models fit to regions for REACT-1 data for sequential rounds; 1 and 2 (yellow), 2 and 3 (blue), 3 and 4 (green), 4 and 5 (pink), 5 and 6 (purple), and 6 and 7a (cyan). Vertical lines show 95% confidence intervals for observed prevalence (black points). Shaded regions show 95% credible intervals for growth models.

Differences in neighbourhood prevalence between rounds 6, 6a, 6b and 7a (smoothed at the level of lower tier local authority, LTLA) reveal variable sub-regional patterns. These mainly show decline especially in North West, North East and Yorkshire and The Humber, but with local areas of growth seen in East and West Midlands (Figure 5, Figure 6). For comparison, unsmoothed LTLA prevalence data are also shown (Figure 7).

**Figure 5.**
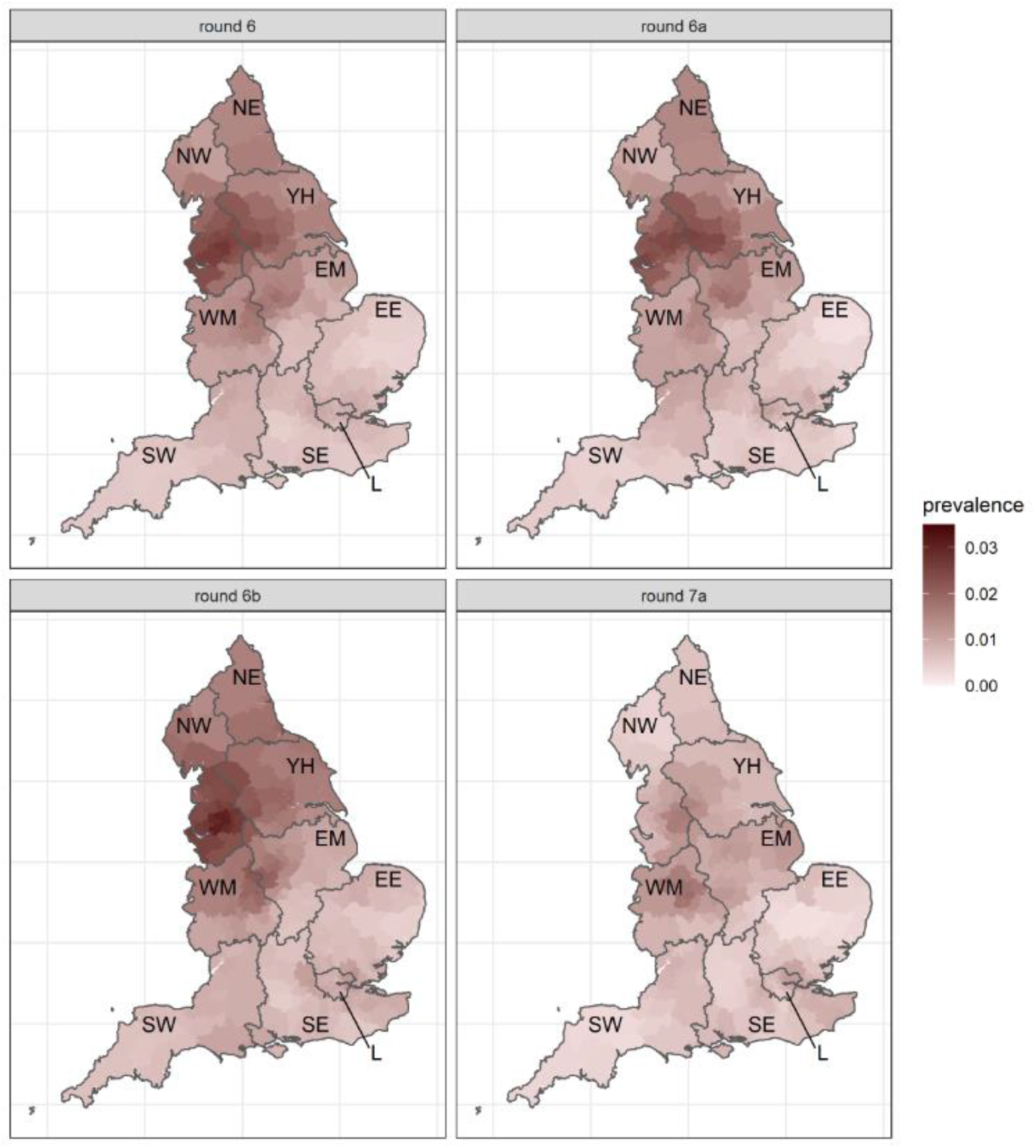
Neighbourhood prevalence for rounds 6, 6a, 6b and 7a. Neighbourhood prevalence calculated from nearest neighbours (the median number of neighbours within 30 km in the study). Average neighbourhood prevalence and difference displayed for individual lower tier local authorities. Regions: NE = North East, NW = North West, YH = Yorkshire and The Humber, EM = East Midlands, WM = West Midlands, EE = East of England, L = London, SE = South East, SW = South West.

**Figure 6.**
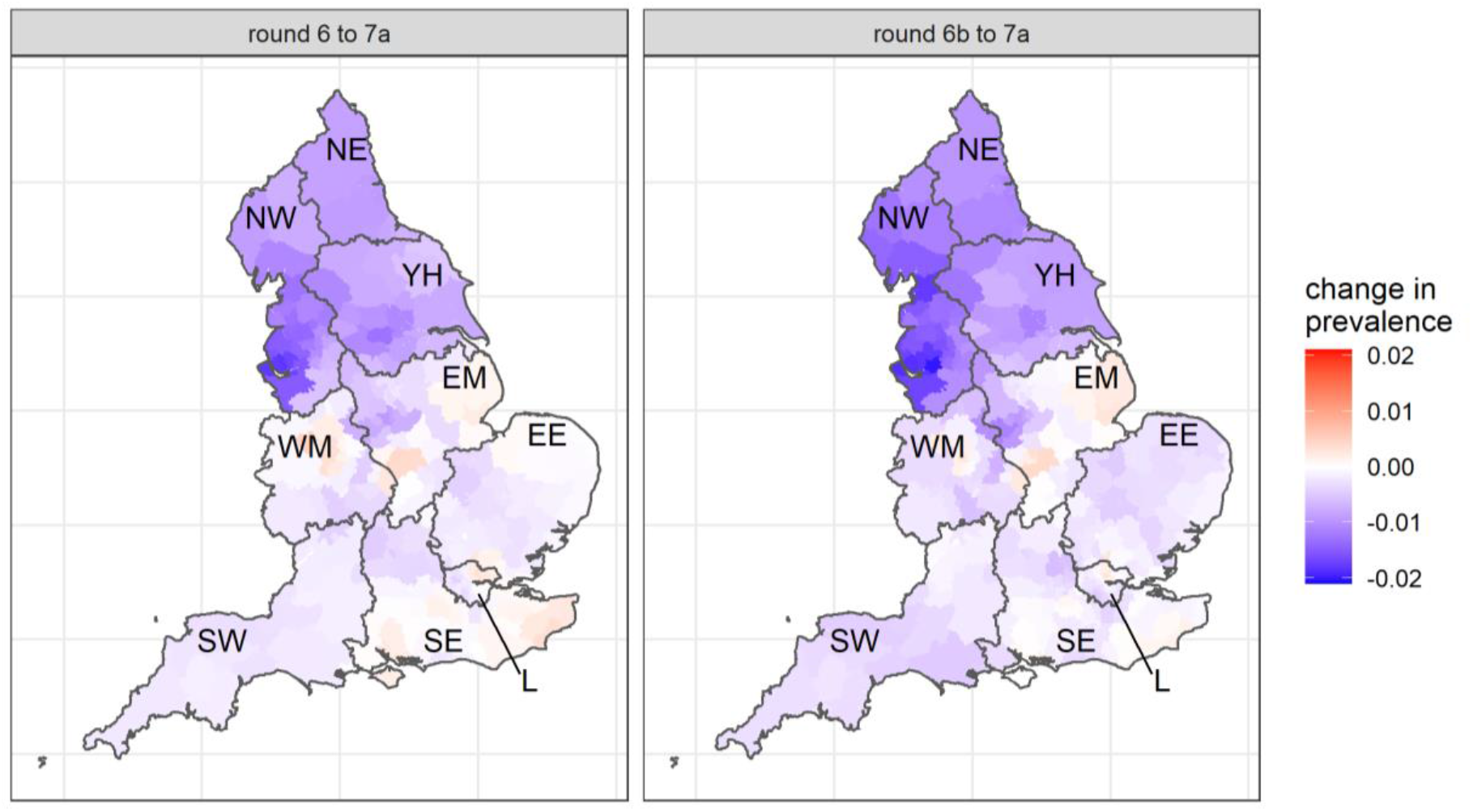
Difference in neighbourhood prevalence at lower tier local authority level between: round 7a and round 6; round 7a and round 6b. Neighbourhood prevalence calculated from nearest neighbours (the median number of neighbours within 30 km in the study). Average neighbourhood prevalence and difference displayed for individual lower tier local authorities. Regions: NE = North East, NW = North West, YH = Yorkshire and The Humber, EM = East Midlands, WM = West Midlands, EE = East of England, L = London, SE = South East, SW = South West.

**Figure 7.**
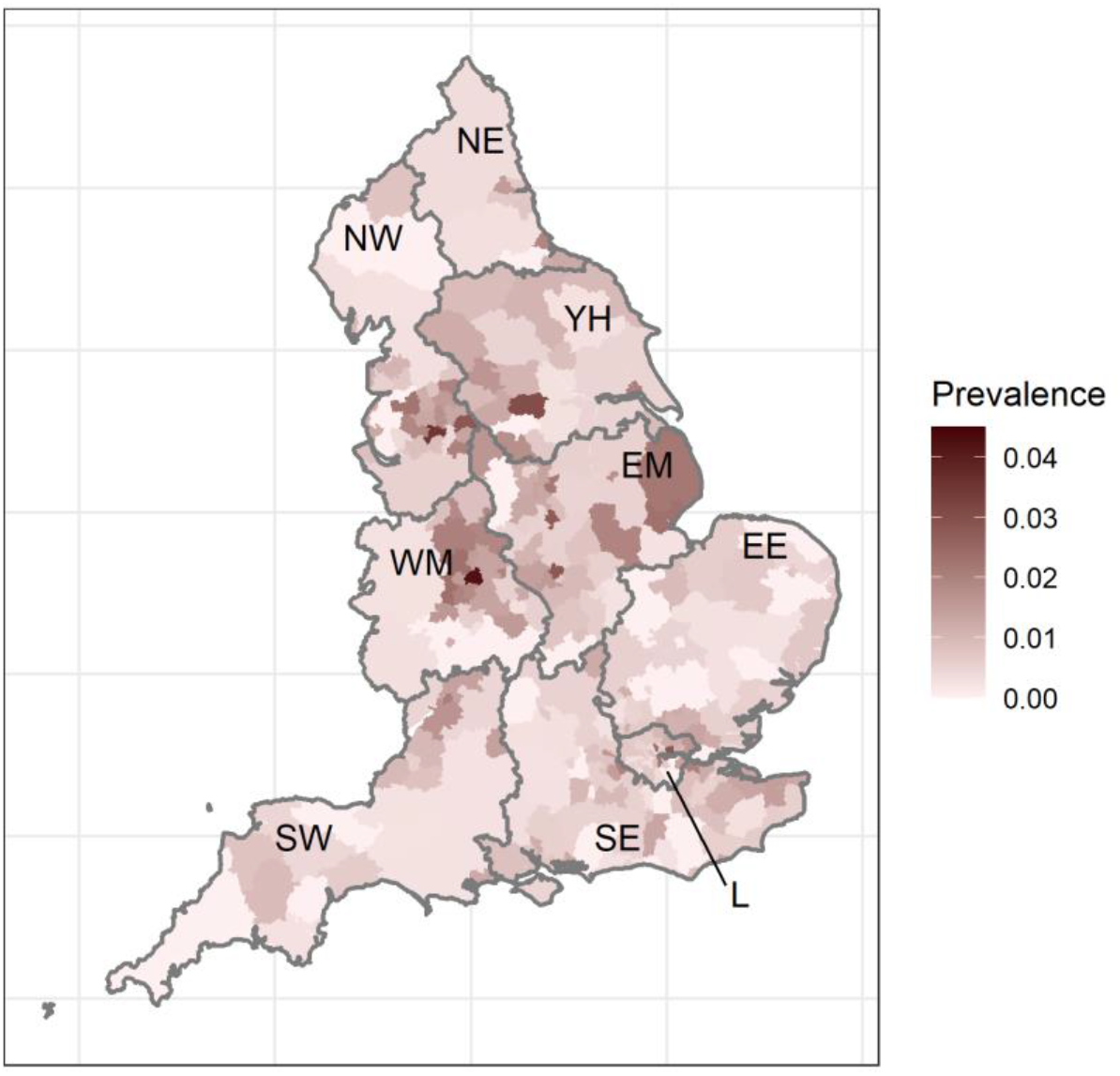
Round 7a unweighted prevalence by lower tier local authority. Regions: NE = North East, NW = North West, YH = Yorkshire and The Humber, EM = East Midlands, WM = West Midlands, EE = East of England, L = London, SE = South East, SW = South West.

In the most recent round 7a data compared with round 6b, there was suggestion of an increase in weighted prevalence in participants aged 5 to 12 years and those aged 13 to 17 years, i.e. among school-aged children, but a decline in all adult age groups (Table 3b, Figure 8). Differences in prevalence between ages at the national level are supported by multivariable logistic regression (Table 5, Figure 9). Our data were also suggestive of age patterns being different between regions (Figure 10). In the North West and Yorkshire and The Humber, among the 18 to 24 year age group, which had the highest prevalence, there was a substantial reduction between rounds 6b and 7a. In contrast, in the West Midlands prevalence in this age group was stable, while it increased in the East Midlands.

**Table 5.**
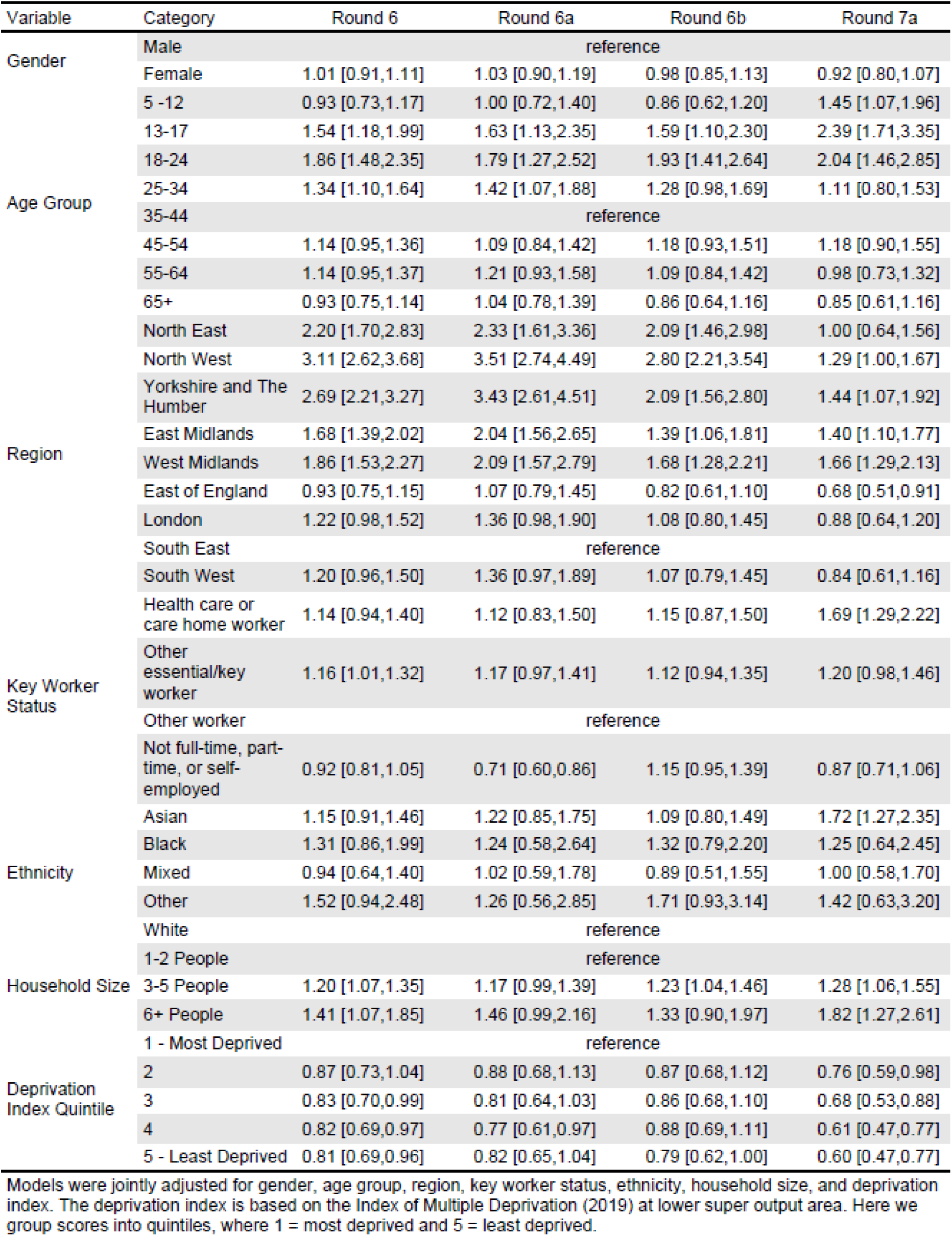
Estimated odds ratios for swab-positivity for rounds 6, 6a, 6b and 7a of the REACT-1 study.

**Figure 8.**
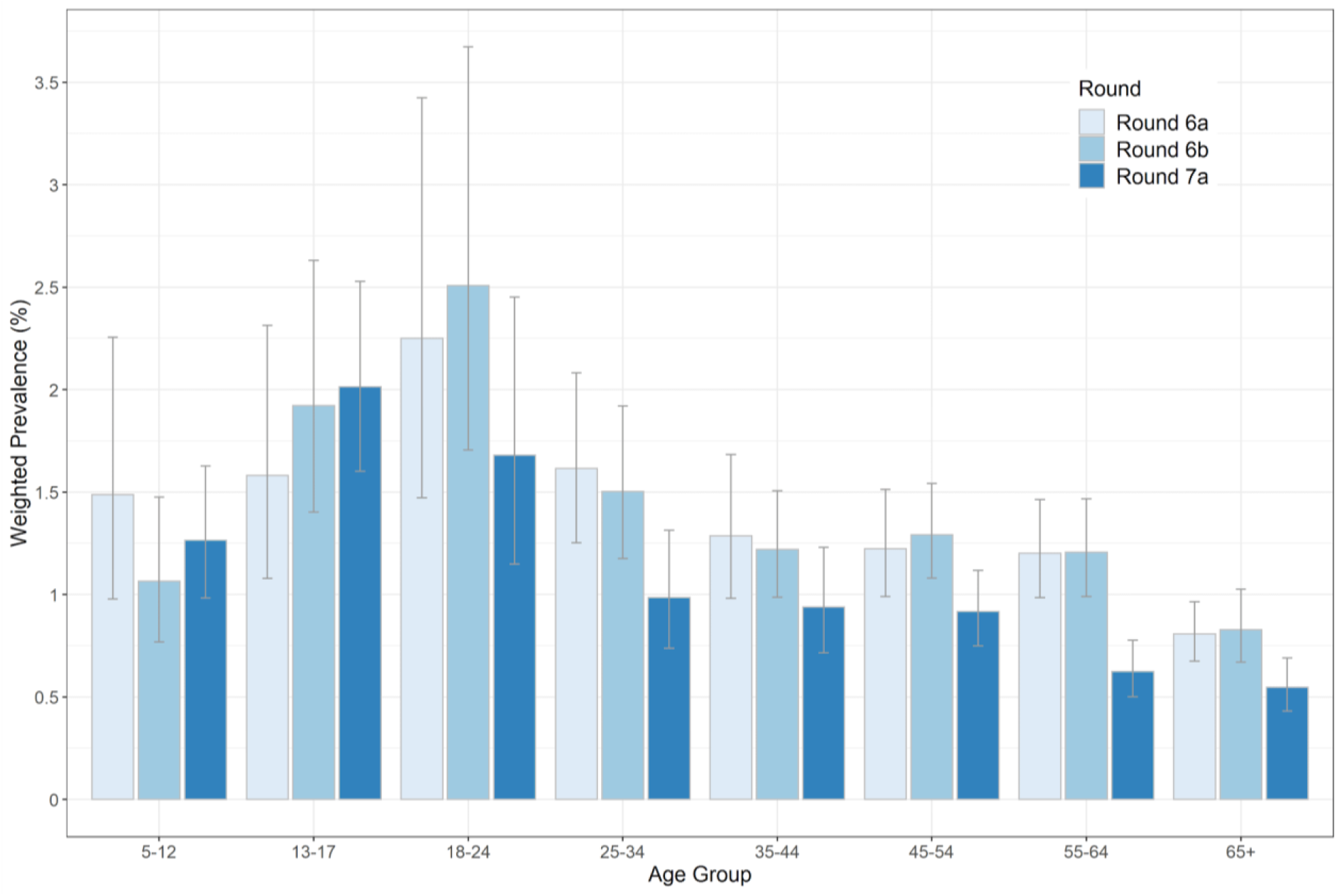
Weighted prevalence of swab positivity by age group for rounds 6a, 6b and 7a. Bars show 95% confidence intervals.

**Figure 9.**
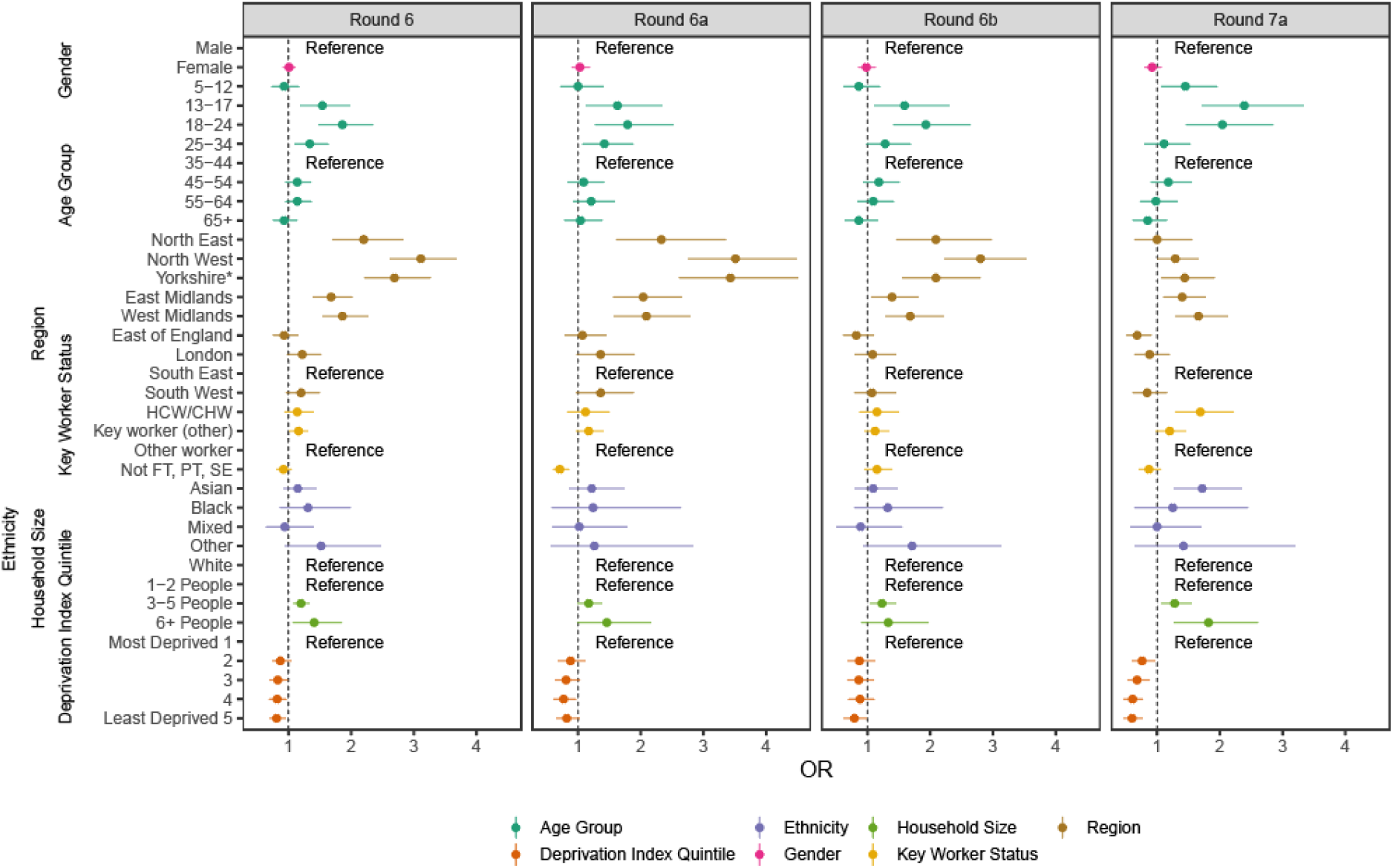
Estimated odds ratios and 95% confidence intervals for jointly adjusted logistic regression model of swab-positivity for rounds 6, 6a (16 to 25 October 2020), 6b (26 October to 2 November 2020), and 7a. Models were jointly adjusted for gender, age group, region, key worker status, ethnicity, household size, and deprivation index. The deprivation index is based on the Index of Multiple Deprivation (2019) at lower super output area. Here we group scores into quintiles, where 1 = most deprived and 5 = least deprived. HCW/CHW = healthcare or care home workers; Not FT, PT, SE = Not full-time, part-time, or self-employed. *Yorkshire and The Humber.

**Figure 10.**
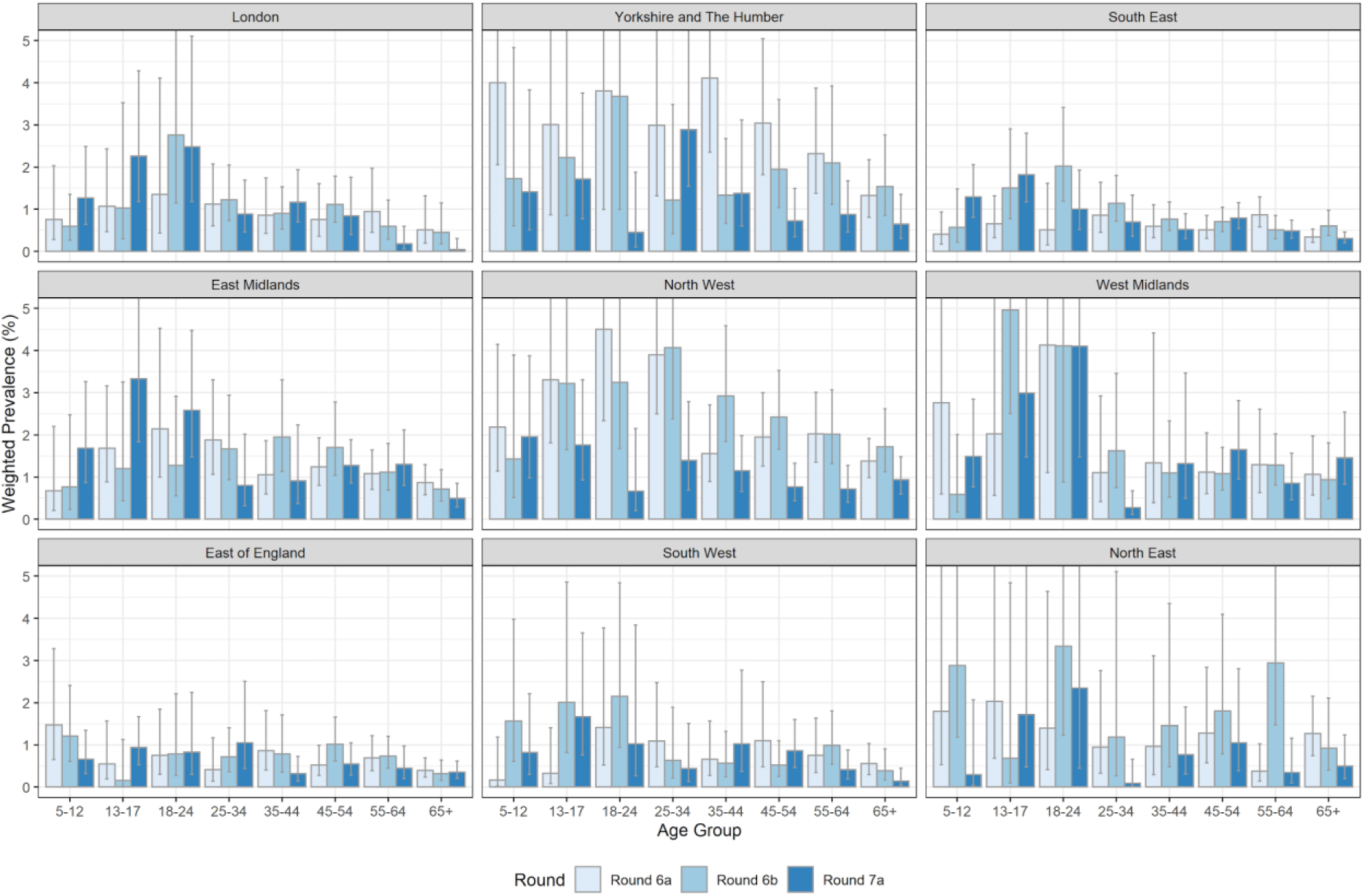
Weighted prevalence of swab positivity by age group and region for rounds 6a, 6b, and 7a. Bars show 95% confidence intervals.

In the most recent round 7a data compared with round 6b, there was no evidence of decline in prevalence among people of Black and Asian ethnicity with an increase in prevalence among people living in the largest households (Table 3b). Further, using an adjusted multivariable logistic regression model, people of Asian ethnicity had increased odds of swab-positivity, 1.72 (1.27, 2.35) during round 7a compared with white people, contrasting with odds of 1.09 (0.80, 1.49) during round 6b (Table 5, Figure 9). Moreover, those living in the most deprived neighbourhoods had higher odds of being swab positive than those living in less deprived neighbourhoods. In addition, healthcare workers and care home workers had 1.69 (1.29, 2.22) increased odds of swab-positivity compared to the employment group “other non-key workers”.

## Discussion

The REACT-1 programme is carrying out population-based surveillance of the SARS-CoV-2 epidemic in England. In this interim report from the seventh round of data collection, we found a reduction in national prevalence of infection by around 30% from the high levels in the latter half of round 6 (26 October to 2 November 2020). The national prevalence has now dropped to ∼1%, a level last seen 6 weeks earlier. This fall in prevalence covers a period of nearly three of the four weeks of the second national lockdown, and is consistent with an observed reduction in the number of daily swab-positive cases recorded in routine surveillance data [5].

The decline in prevalence was especially large in the North where it fell by over 50% in the two regions that had experienced the highest levels in the country during the latter half of round 6 (North West and North East). In contrast, prevalence in London and the Midlands remained almost unchanged. However, the rapid growth of the epidemic seen in London and the South of the country during mid-to late-October [3] was no longer apparent. Our data suggested falls in prevalence across the adult age ranges, including those at older ages who are at the highest risk of severe COVID-19. However, compared with the second half of round 6, prevalence of infection among school aged children (ages 5-12 and 13-17 years) appeared to increase. We note that, in contrast with the first national lockdown in England, schools remained open during this period.

As was the case during the early rounds of REACT-1 [6], during the first half of round 7, swab-positivity was higher among people of Asian ethnicity than white people. We also found higher prevalence among individuals living in the most deprived areas and those living in households with the largest number of people. These findings suggest that social and structural inequalities may be contributing to the transmission of the virus in different communities and that these trends may have been exacerbated during lockdown [7].

We found higher prevalence among healthcare and care home workers during this round compared to other workers which was also the case during the first wave [6], but has been less apparent in our more recent data [1]. This finding indicates possible recent increased transmission of SARS-CoV-2 in clinical and care home settings in England.

We can use our data on prevalence to obtain an estimate of the number of people with detectable virus from a throat and nose swab in England. The national prevalence estimate of 0.96% translates to ∼720,000 infections in England on any one day, with the assumption that sensitivity to detect the virus when present is around 75% [8]. We can also estimate the incidence of infection, assuming an average period of shedding of virus, which we have taken to be ∼10 days. With this assumption, estimated incidence in England is around 72,000 (58,000 to 78,000) infections per day (where the 95% confidence intervals are based on those for the weighted national prevalence estimate). This estimate is down from around 100,000 (90,000 to 104,000) new infections per day at the end of October [1].

Strengths of our study include its size and the use of random community-based sampling to obtain representative estimates of prevalence of SARS-CoV-2 in the population that are not dependent on testing behaviours. Specifically, we are able to detect both symptomatic and asymptomatic infections to provide a more complete picture of the state of the epidemic at any one time than is available from testing only symptomatic individuals. Our study also has a number of limitations. While we do approach a random sample of the population, it is possible that those that respond are, on average, not fully representative of the population as a whole. While we cannot exclude such biases, they are unlikely to materially affect trends in prevalence. We utilise RT-PCR results from self-administered throat and nose swabs which may vary in their ability to detect virus when present, depending on technique of data collection. We correct for sensitivity in our overall estimate of numbers of people infected and, again, this is unlikely to affect trends in prevalence over time. Furthermore, it is possible that changes in sample handling or laboratory procedures could affect swab positivity rates. However, we have well established quality control protocols that should guard against any such systematic differences.

In conclusion, we report a fall in national prevalence and an R number reliably below one during the second national lockdown in England. The largest declines in prevalence since our previous report [1] have been seen in the North of England. These regions include areas that were subject to strict controls on population mixing based on a national tier system that was in place prior to lockdown. After lockdown ends on 2 December 2020, England will return to a tiered local system of restrictions. Despite reductions in national prevalence described here, absolute levels remain high (around 1%). Continued monitoring of the epidemic in the community remains essential until prevalence is reliably suppressed to much lower levels, for example through widespread vaccination.

## Data Availability

The datasets generated or analysed, or both, during this study are not publicly available because of governance restrictions.

## Acknowledgements

SR, CAD acknowledge support: MRC Centre for Global Infectious Disease Analysis, National Institute for Health Research (NIHR) Health Protection Research Unit (HPRU), Wellcome Trust (200861/Z/16/Z, 200187/Z/15/Z), and Centres for Disease Control and Prevention (US, U01CK0005-01-02). GC is supported by an NIHR Professorship. PE is Director of the MRC Centre for Environment and Health (MR/L01341X/1, MR/S019669/1). PE acknowledges support from Health Data Research UK (HDR UK); the NIHR Imperial Biomedical Research Centre; NIHR HPRUs in Chemical and Radiation Threats and Hazards, and Environmental Exposures and Health; the British Heart Foundation Centre for Research Excellence at Imperial College London (RE/18/4/34215); and the UK Dementia Research Institute at Imperial (MC_PC_17114). We thank The Huo Family Foundation for their support of our work on COVID-19.

We thank key collaborators on this work – Ipsos MORI: Kelly Beaver, Sam Clemens, Gary Welch, Nicholas Gilby, and Kelly Ward; Institute of Global Health Innovation at Imperial College: Gianluca Fontana, Dr Hutan Ashrafian, Sutha Satkunarajah and Lenny Naar; North West London Pathology and Public Health England for help in calibration of the laboratory analyses; NHS Digital for access to the NHS register; and the Department of Health and Social Care for logistic support. SR acknowledges helpful discussion with attendees of meetings of the UK Government Office for Science (GO-Science) Scientific Pandemic Influenza – Modelling (SPI-M) committee.

## Tables and Figures

Supporting data to support tables and figures are available [here](https://docs.google.com/spreadsheets/d/1ia14z_YWtWM4AeZnkE0QK7iHbYN0EqJkJeljw7Y7X5s/edit?usp=sharing).

